# Preliminary Analysis of Excess Mortality in India During the Covid-19 Pandemic (Update August 4, 2021)

**DOI:** 10.1101/2021.08.04.21261604

**Authors:** Christopher T. Leffler, Joseph D. Lykins V, Edward Yang

## Abstract

**Background:** As both testing for SARS Cov-2 and death registrations are incomplete or not yet available in many countries, the full impact of the Covid-19 pandemic is currently unknown in many world regions.

**Methods:** We studied the Covid-19 and all-cause mortality in 18 Indian states (combined population of 1.26 billion) with available all-cause mortality data during the pandemic for the entire state or for large cities: Gujarat, Karnataka, Kerala, Maharashtra, Tamil Nadu, West Bengal, Delhi, Madhya Pradesh, Andhra Pradesh, Telangana, Assam, Bihar, Odisha, Haryana, Rajasthan, Himachal Pradesh, Punjab, and Uttar Pradesh. Excess mortality was calculated by comparison with available data from years 2015-2019. The known Covid-19 deaths reported by the Johns Hopkins University Center for Systems Science and Engineering for a state were assumed to be accurate, unless excess mortality data suggested a higher toll during the pandemic. Data from Uttar Pradesh were not included in the final model due to anomalies.

**Results:** In several regions, fewer deaths were registered in 2020 than expected. The excess mortality in Mumbai (in Maharashtra) in 2020 was 137.0 / 100K. Areas in Tamil Nadu, Kolkata (in West Bengal), Delhi, Madhya Pradesh, Karnataka, Haryana, and Andhra Pradesh saw spikes in mortality in the spring of 2021.

**Conclusions:** The pandemic-related mortality through June 30, 2021 in 17 Indian states was estimated to be 132.9 to 194.4 per 100,000 population. If these rates apply to India as a whole, then between 1.80 to 2.63 million people may have perished in India as a result of the Covid-19 pandemic by June 30, 2021. This per-capita mortality rate is similar to that in the United States and many other regions.

## Introduction

As both testing for SARS Cov-2 and death registrations are incomplete or not yet available in many countries, the full impact of the Covid-19 pandemic is unknown in many world regions. One approach to assess the impact of the evolving pandemic is to examine excess mortality from all causes. When all-cause mortality increases during the pandemic, this is assumed to be a direct result of infection with the Sars Cov-2 virus, or indirect effects from health system overload or social responses to the pandemic.

For many countries, national tallies of mortality during portions of the pandemic have already become available. In India, regional government websites and data journalists are publishing up-to-date mortality figures for an ever-increasing number of cities and states. We sought to integrate these data to estimate the impact of the Covid-19 pandemic in India as a whole. We understand that the picture might change as the pandemic proceeds, and as more data become available.

## Methods

We used the publicly available mortality figures published by regional governments (Kerala, Odisha, Karnataka, and Tamil Nadu) and by data journalists in India, often obtained from Right-to-Information (RTI) requests (Hindustan Times 2021; Ramani “Hindu” 2021; Ramani “docs” 2021; Ramani “Rajasthan” 2021; Ramani “Haryana” 2021; Ramani “Punjab”; Scroll 2021; Nagar 2021; The Hindu 2020; Khanna 2020; Saikia 2021; Radhakrishnan 2021, Rukmini 2021; Rukmini “Andhra” 2021). Much of these data are stored on the Local Mortality GitHub websites maintained by Ariel Karlinsky, Dmitry Kobak, and colleagues (Karlinsky 2021; Karlinsky & Kobak 2021) or on the Development Data Lab website (devdatalab).

We assumed that the mortality rate related to the Covid-19 pandemic in a given region was equivalent to the mortality reported by the Johns Hopkins University Center for Systems Science and Engineering (CSSE) (based on positive viral testing and clinical symptoms) (2021), unless the excess mortality data suggested a higher toll. When data for a given region and time period were conflicting or contradictory, we reported a range of estimates to reflect the uncertainty.

All of these publicly available regional-level mortality data contain no individually-identifiable information. The study was approved by the Office of Research Subjects Protection of Virginia Commonwealth University.

For Chhattisgarh, mortality data from the online portal of the state CRS have been released by the Development Data Lab, and we included these data in the appendix. However, as the baseline data for this online portal appear to be only about one tenth complete, as compared with the national vital statistics registry, we deemed the Chhattisgarh data too unreliable to include in the model.

For Uttar Pradesh, the raw mortality data obtained from a Right-to-Information request contained anomalies, such as multiple districts with zero deaths for numerous months. Therefore, the Uttar Pradesh data were analyzed, but were not included in the top-line model.

For the urban portions of 25 districts in Madhya Pradesh, the numbers of funerals in April 2021 have been tabulated (Datta 2021). For Gujarat and the urban portions of 25 districts in Madhya Pradesh, mortality from entire year(s) before 2020 was available (Vital Statistics reports). Therefore, to estimate excess mortality for portions of 2021, it was necessary to assume that mortality was evenly distributed throughout the year.

We calculated excess mortality in a region by comparing the mortality for a given time period in 2020 or 2021 with the value expected based on the years 2015 to 2019. If data from more than one year before 2019 was available, the expected value was calculated by creating a trend line for mortality by linear regression for the years 2015 to 2019, and carrying this trend one year (for 2020) or two years (for 2021) into the future. Carrying the trend line two years into the future for 2021 yielded conservative estimates of excess deaths.

For some states, reported mortality from the state government websites or right-to-information (RTI) requests was only available for 2018 and 2019, which was too short a period to generate a robust trend line. Moreover, the numbers of deaths from the state sources did not match the central government figures exactly, because the state information systems did not capture all of the registered deaths. In these cases, the vital statistics reports for India were used to generate a trend line for expected deaths, using the data from 2015 to 2019. The expected number of deaths was scaled up or down by multiplying the 2015 to 2019 trend line by the ratio of deaths in the state and federal systems for 2018 and 2019. For instance, if the state website average mortality for 2018 and 2019 was 97% of the figures for 2018 and 2019 in the federal reports, the trend line was multiplied by 0.97. This method was used to scale the trend line for Delhi, Bengaluru, Mumbai, Nagpur, Ahmedabad (for 2020), Madhya Pradesh, Tamil Nadu, for 6 city hospitals in Tamil Nadu, and for Madurai district.

Completeness of death registrations has been estimated in the vital statistics reports for each state by comparison with the Sample Registration System (Table S1). For years in which the completeness was less than 100%, the total number of deaths for each year 2015 to 2019 was determined by dividing the death registrations by the completeness fraction. Unlike the trend line for unadjusted death registrations, the trend line for the adjusted registrations did decrease over time for some states. In order to ensure the estimated excess deaths were conservative, the expected deaths for 2020 and 2021 were the maximum of the 2019 value and the value predicted by the trend line. The completeness fraction for 2020 and 2021 was extrapolated by linear regression from the 2015 to 2019 completeness fraction. Once again, in order to be conservative, the maximum of the 2019 value and the linear extrapolation was used.

The national mortality rate was estimated by summing the estimated pandemic-related deaths for the states analyzed and then dividing by the population of these states. The population of Indian states was taken from the Hopkins mortality dataset. Raw data are tabulated in the appendix (Table S1).

## Results

We studied 17 states for which excess mortality data were available for at least a portion of the state: Gujarat, Karnataka, Kerala, Maharashtra, Tamil Nadu, West Bengal, Delhi, Madhya Pradesh, Andhra Pradesh, Telangana, Assam, Bihar, Odisha, Rajasthan, Haryana, Punjab and Uttar Pradesh. These 17 states have a combined population of 1.26 billion people (Table 1).

**Table 1.** Covid-19 Deaths Based on Viral Testing and Clinical Symptoms, as Tabulated by Johns Hopkins University.

| Region. | Population | Reported Covid Deaths in 2020 | Deaths/100K in 2020 | Reported Covid deaths by June 30, 2021. | Deaths/100K in 2021 (as of June 30) |
| --- | --- | --- | --- | --- | --- |
| Delhi | 18,710,920 | 10,523 | 56.240 | 24,971 | 77.217 |
| Karnataka | 67,562,700 | 12,081 | 17.881 | 34,929 | 33.817 |
| Maharashtra | 123,144,200 | 49,463 | 40.167 | 121,804 | 58.745 |
| Gujarat | 63,872,400 | 4,302 | 6.735 | 10,056 | 9.009 |
| Tamil Nadu | 77,841,270 | 12,109 | 15.556 | 32,506 | 26.203 |
| West Bengal | 99,609,300 | 9,683 | 9.721 | 17,679 | 8.027 |
| Kerala state | 35,699,440 | 3,042 | 8.521 | 13,093 | 36.676 |
| Madhya Pradesh | 85,358,970 | 3,595 | 4.212 | 8,954 | 6.278 |
| Andhra Pradesh | 53,903,393 | 7,104 | 13.179 | 12,671 | 10.328 |
| Telangana | 39,362,732 | 1,541 | 3.915 | 3,651 | 5.360 |
| Assam State | 35,607,039 | 1,043 | 2.929 | 4,509 | 9.734 |
| Bihar | 124,799,930 | 1,393 | 1.116 | 9,584 | 6.563 |
| Uttar Pradesh | 237,882,725 | 8,352 | 3.511 | 22,577 | 5.980 |
| Rajasthan | 81,032,689 | 2,689 | 3.318 | 8,918 | 7.687 |
| Odisha | 46,356,334 | 1,871 | 4.036 | 3,970 | 4.528 |
| Haryana | 28,204,692 | 2,899 | 10.278 | 9,417 | 23.110 |
| Punjab | 30,141,373 | 5,331 | 17.687 | 16,033 | 35.506 |
| Himachal Pradesh | 7,451,955 | 931 | 12.493 | 3,477 | 34.166 |
| Total | 1,256,542,062 | 137,952 | 10.979 | 358,799 | 17.576 |

Data from June 30, 2021. This table lists absolute and per-capita Covid-19 deaths, rather than excess deaths.

### Reported Covid-19 Mortality

The mortality related to Covid-19, based on viral testing and the clinical picture, as tabulated by Johns Hopkins, was reasonably low: 11.0 / 100K in 2020, and 17.6 / 100K in 2021, for a combined total of 28.6 / 100K for the pandemic, as of June 30, 2021 (Table 1). There was some variation, with lower mortality rates in Gujarat, West Bengal, Telangana, Assam, Odisha, Uttar Pradesh, Rajasthan, and Bihar and higher mortality rates in Maharashtra, Delhi, and Punjab (Table 1).

### Excess Mortality

Excess mortality data was available for regions of 18 states, including 17 states in 2020 (Table 2), and 17 states in 2021 (Table 3). The best available estimates of the mortality range, whether based on reported Covid-19 deaths, or on excess mortality, are tabulated in Table 4. Table 5 presents the top-line model to estimate excess mortality during the pandemic in India.

**Table 2.** Excess mortality in India from Jan. 1, 2020 to Dec. 31, 2020.

| State | Region. | Population | Period in 2020 | Excess deaths | Deaths/100K |
| --- | --- | --- | --- | --- | --- |
| Dehli |  | 18,710,920 | Apr-May | -9,702 | -51.852 |
| Bihar |  | 124,799,930 | Entire year | 134,244 | 107.567 |
| Karnataka | Entire state | 67,562,700 | Entire year | 12,627 | 18.689 |
|  | Bengaluru | 8,443,675 | Jan-Jul | 9,908 | 117.342 |
| Maharashtra | State less 6 districts | 102,598,050 | Entire year | 103,152 | 100.540 |
|  | Mumbai | 12,875,213 | Entire year | 17,641 | 137.018 |
|  | Nagpur | 2,405,000 | Apr-Dec | 4,144 | 172.308 |
| Gujarat | Gujarat | 63,872,400 | Jan-Nov | -70,333 | -110.115 |
|  | Ahmedabad | 8,059,000 | Apr to May | 5,276 | 65.466 |
| Tamil Nadu | Tamil Nadu | 77,841,270 | Entire year | 72,052 | 92.563 |
|  | Chennai | 7,088,000 | Entire year | 5,921 | 83.540 |
|  | 6 cities | 5,980,370 | Jan-May | -1,032 | -17.261 |
|  | Madurai dist. | 3,038,250 | Entire year | 2,018 | 66.412 |
| West Bengal | Entire state | 99,609,300 | Entire year | 53,355 | 53.564 |
|  | Kolkata | 4,496,694 | Entire year | 2,075 | 46.145 |
| Telangana | Hyderabad | 9,482,000 | Entire year | 8,359 | 88.157 |
| Haryana |  | 28,204,692 | Entire year | 11,087 | 39.311 |
| Kerala |  | 35,699,440 | Entire year | -27,367 | -76.659 |
| Madhya Pradesh |  | 85,358,970 | Entire year | -28,598 | -33.503 |
| Andhra Pradesh |  | 53,903,393 | Entire year | 65,493 | 121.501 |
| Assam State |  | 35,607,039 | Entire year | 15,246 | 42.819 |
| Uttar Pradesh |  | 237,882,725 | Entire year | -37,468 | -15.751 |
| Rajasthan |  | 81,032,689 | Entire year | 6,629 | 8.181 |
| Punjab |  | 30,141,373 | Entire year | -11,415 | -37.873 |
| Himachal Pradesh |  | 7,451,955 | Entire year | 291 | 3.902 |

The 6 government hospitals in Tamil Nadu studied by Arappor Iyakkam from Jan-May 2020 were in Madurai, Coimbatore, Trichy, Vellore, Karur, and Tirupur. Maharashtra excess mortality did not include 6 districts: Pune, Satara, Sangli, Yavatmal, Sindhudurg and Gondiya (Ramani).

**Table 3.**
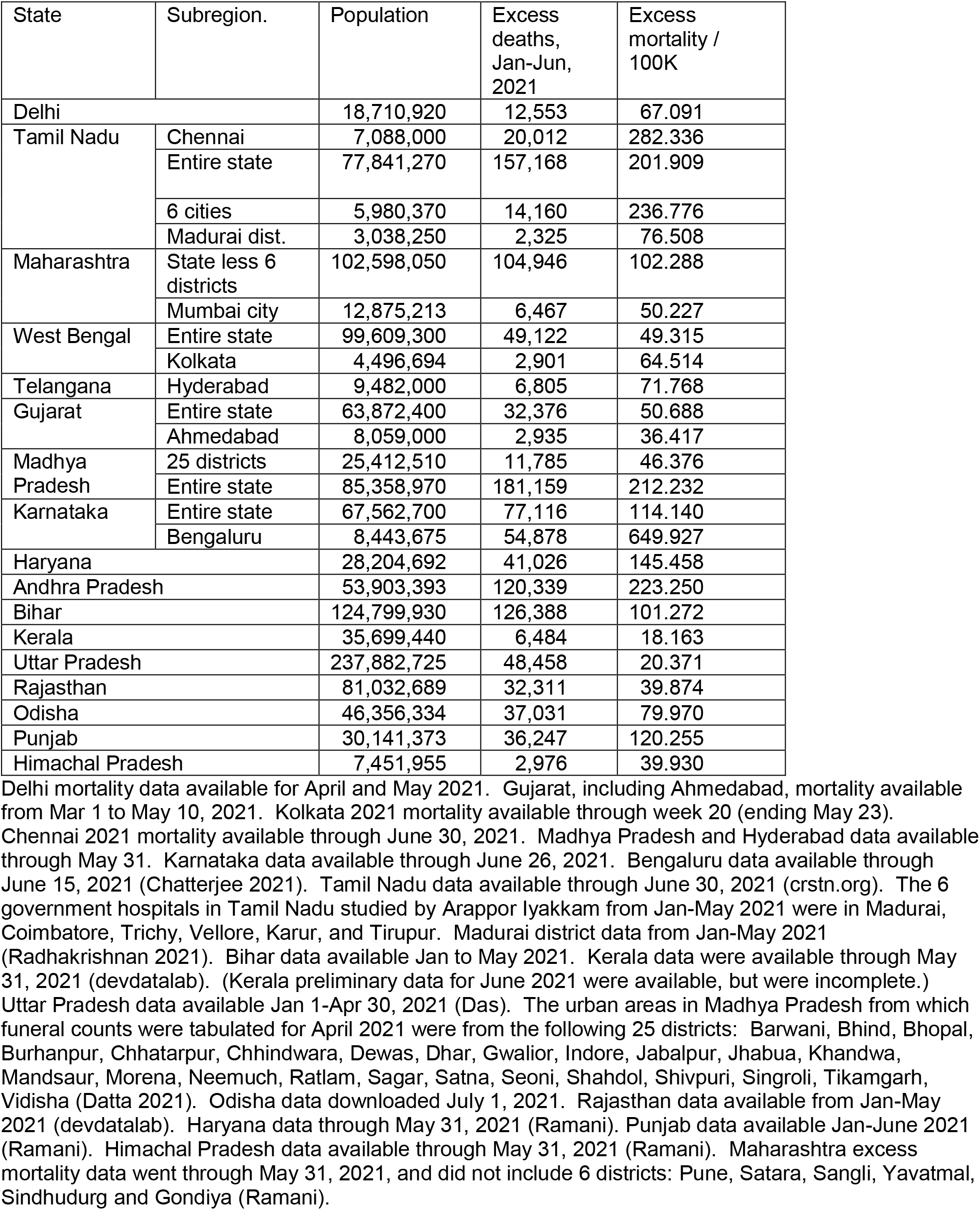
Excess mortality in India, early 2021.

**Table 4.** Estimated Covid-19 Deaths in India in 2020 and early 2021.

| State | Population | Type | Data Source(s) | 2020 |  | 2021, by June 30 |  |
| --- | --- | --- | --- | --- | --- | --- | --- |
|  |  |  |  | Deaths/100K | Deaths | Deaths/100K | Deaths |
| Delhi | 18,710,920 | -- | Hopkins | 56.240 | 10,523 | 77.217 | 14,448 |
| Karnataka | 67,562,700 | Low | Karnataka | 18.689 | 12,627 | 114.140 | 77,116 |
|  |  | High | Bengaluru | 117.342 | 79,279 |  |  |
| Maharashtra | 123,144,200 | Low | State,Mumbai | 100.540 | 123,809 | 50.227 | 61,852 |
|  |  | High | Nagpur,State | 172.308 | 212,187 | 102.288 | 125,962 |
| Gujarat | 63,872,400 | Low | Hopkins, Gujarat | 6.735 | 4,302 | 50.688 | 32,376 |
|  |  | High | Ahmedabad | 65.466 | 41,815 |  |  |
| Tamil Nadu | 77,841,270 | Low | Madurai | 66.412 | 51,696 | 76.508 | 59,555 |
|  |  | High | Tamil Nadu, Chennai | 92.563 | 72,052 | 282.336 | 219,774 |
| West Bengal | 99,609,300 | Low | Kolkata, | 46.145 | 45,965 | 49.315 | 49,122 |
|  |  | High | Entire state | 53.564 | 53,355 | 64.514 | 64,262 |
| Telangana | 39,362,732 | -- | Hyderabad | 88.157 | 34,701 | 71.768 | 28,250 |
| Kerala | 35,699,440 | -- | Hopkins | 8.521 | 3,042 | 36.676 | 13,093 |
| Madhya Pradesh | 85,358,970 | Low | Hopkins, 25 Districts | 4.212 | 3,595 | 46.376 | 39,586 |
|  |  | High | Entire state |  |  | 212.232 | 181,159 |
| Andhra Pradesh | 53,903,393 | -- | Entire state | 121.501 | 65,493 | 223.250 | 120,339 |
| Assam | 35,607,039 | -- | Entire state | 42.819 | 15,246 | -- | -- |
| Bihar | 124,799,930 | -- | Entire state | 107.567 | 134,244 | 101.272 | 126,388 |
| Rajasthan | 81,032,689 | -- | Entire state | 8.181 | 6,629 | 39.874 | 32,311 |
| Haryana | 28,204,692 | -- | Entire state | 39.311 | 11,087 | 145.458 | 41,026 |
| Odisha | 46,356,334 | -- | Entire state | -- | -- | 79.970 | 37,071 |
| Punjab | 30,141,373 | -- | Hopkins, Punjab | 17.687 | 5,331 | 120.255 | 36,247 |
| Himachal Pradesh | 7,451,955 | -- | Hopkins, Himachal Pradesh | 12.493 | 931 | 39.930 | 2,976 |
| Uttar Pradesh | 237,882,725 | -- | Entire state | 3.511 | 8,352 | 20.371 | 48,458 |
| Total, excluding Uttar Pradesh | 972,303,003 (2020); 983,052,298 (2021) | Low |  | 54.430 | 529,221 | 78.506 | 771,756 |
|  |  | High |  | 77.086 | 749,510 | 117.267 | 1,152,798 |
| Total. | 1,210,185,728 (2020); 1,220,935,023 (2021) | Low |  | 44.421 | 537,573 | 67.179 | 820,214 |
|  |  | High |  | 62.624 | 757,862 | 93.388 | 1,201,256 |

All estimates based on excess deaths, except 2020 Delhi, Kerala, and Punjab data, which are based on known Covid-19 mortality as reported by Johns Hopkins University, and Gujarat, West Bengal, Madhya Pradesh, and Tamil Nadu, which included the Hopkins data as the low end of the uncertainty range.

**Table 5.** Estimated Excess Mortality in India.

|  | Excess Mortality (per 100,000) |  | Excess deaths |  |
| --- | --- | --- | --- | --- |
|  | Low | High | Low | High |
| 2020 | 54.430 | 77.086 | 736,243 | 1,042,698 |
| 2021 | 78.506 | 117.267 | 1,061,905 | 1,586,203 |
| Both years | 132.936 | 194.353 | 1,798,149 | 2,628,901 |

Assumes population of India of 1,352,642,280. Per-capita excess mortality based on all states analyzed except Uttar Pradesh.

For Kerala, Delhi, Madhya Pradesh, Punjab, and Uttar Pradesh, the excess mortality based on registered deaths was actually negative for 2020. This may be because fewer people were willing to register deaths during lockdown, or because fewer people died from accidents and other causes during lockdown (Table 1).

At the other extreme, the excess mortality in Mumbai (in Maharashtra) was 137.0 / 100K in 2020 (Tables 2 and 4). Similarly, Bengaluru in Karnataka had an excess mortality of 117.3 / 100K in the first 7 months of 2020 (Tables 2 and 4).

For 2020, intermediate levels of excess mortality were seen for Kolkata in West Bengal (46.1 / 100K), Chennai in Tamil Nadu (83.5 / 100K), and Hyderabad in Telangana (88.2 / 100K) (Tables 2, 4).

For 2021, a significant peak in all-cause mortality was seen for March through June for Chennai in Tamil Nadu, Kolkata in West Bengal, Delhi, Madhya Pradesh, Haryana, Punjab, and Andhra Pradesh (Tables 3, 4). These findings correspond with news reports of increasing severity of the pandemic in India. During the first half of 2021, the excess mortality was 64.5 / 100K for Kolkata, 67.1 / 100K for Delhi, 120.3 / 100K for Punjab, 145.5 / 100K in Haryana, 176.8 / 100K for Madhya Pradesh, 197.9 / 100K for Andhra Pradesh, and 282.3 / 100K for Chennai in Tamil Nadu (Tables 3, 4).

### Integrated Model of Covid-19-related Mortality

A model of pandemic-related mortality which integrates the available data is shown in Table 4. Generally, the excess mortality exceeded the official Covid-19 mortality figures, and was therefore taken as the pandemic-related mortality. However, the Hopkins data on reported Covid-19 deaths were used for Kerala, Delhi, Himachal Pradesh, and Punjab in 2020, and for the low end of the uncertainty range for several regions.

Despite the wide uncertainty ranges for several states and time periods, the overall uncertainty range was more narrow. In the primary model, data from Uttar Pradesh were excluded because of identified anomalies. For 2020, the pandemic-related mortality for 16 states with a population of 972,303,003 million was estimated to be 54.4 to 77.1 / 100K (Table 4). For 2021, through June 30, the pandemic-related mortality for 16 states with a population of 983,052,298 million was estimated to be 78.5 to 117.3 / 100K (Tables 4,5).

Summing these estimates for 2020 and 2021, we estimate the pandemic-related mortality to be: 132.9 to 194.4 per 100,000 population for the entire pandemic (through June 30, 2021, Table 5). Assuming a population of India of 1,352,642,280, these rates correspond with a mortality range of 1.80 to 2.63 million people perishing during the pandemic in India from Covid-19 by June 30, 2021.

If the data from Uttar Pradesh are included, then the estimated pandemic-related mortality was: 111.6 to 161.0 per 100,000 population for the entire pandemic (through June 30, 2021), corresponding with a mortality range of 1.51 to 2.18 million people perishing during the pandemic in India from Covid-19.

## Discussion

This analysis of excess mortality found that between 1.80 to 2.63 million people may have perished in India as a result of the Covid-19 pandemic, as of June 30, 2021. It should be noted that the estimated per-capita mortality of 133 to 194 per 100,000 in India is similar to that of the United States and many other regions.

Data from Uttar Pradesh contained anomalies and the estimated excess mortality was lower than in other regions. However, if the Uttar Pradesh data are included in the model, the estimated pandemic-related mortality in India through June 30, 2021 was 1.51 to 2.18 million people.

This mortality level is well above the reported Covid-19 mortality of 398,454 in India as of June 30, 2021 (Hopkins). The IHME at the University of Washington currently estimates that the excess mortality in India was 1.14 million persons on June 30, 2021 (IHME 2021a). It should be noted, however, that the IHME’s original model did not look directly at all-cause mortality in India. Rather, the IHME model extrapolated Indian all-cause mortality based on factors such as test positivity rates in India, and all-cause mortality data from other countries, such as Mexico, Brazil, and the United States (IHME 2021b). Our analysis was based on actual counts of mortality in India, and therefore was a more direct approach to estimation.

Our method of determining the expected baseline, by making a projection using linear regression one year (for 2020) or two years (for 2021) into the future, was more conservative than simply using the baseline average mortality, or projecting just one year forward (even for 2021). In other words, our analysis used the accepted Covid mortality, unless there was very compelling excess mortality data to reject the official numbers. Thus, our estimates of pandemic-related mortality for various regions are more conservative (i.e. lower) than some other studies and news reports. For instance, Deshmukh and colleagues estimated excess mortality in India of 3.2 million based on Civil Registration System data from 5 states and 5 cities (Deshmukh 2021). Anand and colleagues estimated excess deaths of 3.4 million based on death registrations from seven states, including the somewhat problematical data Chhattisgarh data (Anand 2021). If the number of death registrations increased by a fixed amount every year between 2015 and 2021 in a given state, our study would have reported no excess deaths, while the studies of Deshmukh and Anand would have attributed each annual step increase to the pandemic. Their approach may ultimately prove to generate more accurate estimates, but in the face of uncertainty, we elected to take a more conservative approach. Our definition of the baseline by linear regression was used previously in the World Mortality Dataset (Karlinsky 2021).

Based on mortality data from the online Health Management Information System of the Ministry of Health and Family Welfare, Deshmukh estimated excess deaths in India of 2.7 million through June 2021 (Deshmukh 2021).

Survey data can supplement mortality estimates from death registrations. Based on the consumer pyramid household survey (CPHS) produced by the Center for the Monitoring of the Indian Economy (CMIE), Anand estimated excess mortality in India during the pandemic of 4.9 million persons (Anand 2021). Based on a national telephone survey conducted by Cvoter India OmniBus, Deshmukh estimated 3.1 to 3.4 million excess deaths through June 2021 (Deshmukh 2021). One limitation of this survey is that it included deaths outside the immediate household (Deshmukh 2021).

Seroprevalence data can also help to assess the extent of infection in a population. Based on application of international age-specific infection fatality rates to Indian demography and seroprevalence, Anand estimated the Covid-19 related mortality in India to be 4.0 million (Anand 2021). One limitation of this approach is that the infection fatality rates may differ from country to country, depending on the medical system and host factors.

Guilmoto analyzed Covid-19 mortality in several well-defined Indian populations: deaths in Kerala, elected representatives, Indian Railways personnel, and teachers in Karnataka (Guilmoto 2021). Application of the age-specific mortality rates to the Indian population yielded a mortality estimate of 2.2 million persons by late May 2021 (Guilmoto 2021).

Our analysis has a number of limitations. The data analyzed are still incomplete for many regions and times. There may be delays in registering deaths. In addition, available data obtained from regional government websites, central government compilations, and by reporters through RTI requests are not in complete agreement. Some of the data upon which the model is based may simply be wrong, because early reports may contain errors. All-cause mortality may be higher not only due to infection with the Sars Cov-2 virus, but also because of health system overload, delays in patients entering the health system for other conditions (Wu 2020, Aldujeli 2020), or social changes, such as lockdowns. A number of countries, such as Australia and New Zealand, have experienced lower than normal mortality during the pandemic (Karlinsky “World Mortality”). Lower mortality rates may occur because there are fewer accidents or homicides, etc. On the other hand, other factors may lead to higher mortality rates during lockdowns in some regions.

Time will tell what the true death toll has been, as the early data are confirmed, additional regions provide more complete mortality data, and data from diverse sources are reconciled. Additional surveys and seroprevalence data can supplement the estimates from death registrations.

## Data Availability

Data are in the appendix of the article.

## Acknowledgments

The author would like to acknowledge the data journalists, academicians, and nonprofits in India who have brought these data to light, such as Rukmini S. (@Rukmini), Chinmay Tumbe (@ChinmayTumbe), Vignesh Radhakrishnan (@VigneshJourno), Srinivasan Ramani (@vrsrini), Deepak Patel (@deepakpatel_91), Murad Banaji (@muradbanaji), Sumitra Debroy (@debroysumitra), Dhanya Rajendran (@dhanyarajendran), Shiba Kurian (@shiba_kurian), and Arappor Iyakkam. In addition, we acknowledge the important work of Ariel Karlinsky and colleagues, and the Development Data Lab for preparing many of the mortality databases we used. However, all errors in the paper are the responsibility of the authors.

## Appendix

**Table S1.** Time Series of Mortality from Selected Regions in India.

| Region | Year | Period | Deaths | Registration Completeness | Reference |
| --- | --- | --- | --- | --- | --- |
| Andhra Pradesh | 2015 | All | 310,640 | 0.849 | Vital Statistics |
| Andhra Pradesh | 2016 | All | 313,285 | 0.887 | Vital Statistics |
| Andhra Pradesh | 2017 | All | 355,546 | 0.944 | Vital Statistics |
| Andhra Pradesh | 2018 | All | 375,777 | 1.000 | Vital Statistics |
| Andhra Pradesh | 2019 | All | 401,472 | 1.000 | Vital Statistics |
| Andhra Pradesh | 2018 | All | 333,275 |  | Rukmini "AP" |
| Andhra Pradesh | 2019 | All | 363,414 |  | Rukmini "AP" |
| Andhra Pradesh | 2020 | All | 428,907 |  | Rukmini "AP" |
| Andhra Pradesh | 2018 | Months 1-5 | 140,102 |  | Rukmini "AP" |
| Andhra Pradesh | 2019 | Months 1-5 | 147,662 |  | Rukmini "AP" |
| Andhra Pradesh | 2020 | Months 1-5 | 134,632 |  | Rukmini "AP" |
| Andhra Pradesh | 2021 | Months 1-5 | 272,910 |  | Rukmini "AP" |
| Assam State | 2015 | All | 116,778 | 0.511 | Vital Statistics |
| Assam State | 2016 | All | 130,414 | 0.598 | Vital Statistics |
| Assam State | 2017 | All | 141,012 | 0.659 | Vital Statistics |
| Assam State | 2018 | All | 142,605 | 0.669 | Saikia, Vital St. |
| Assam State | 2019 | All | 163,057 | 0.740 | Saikia, Vital St. |
| Assam State | 2020 | All | 187,085 |  | Saikia |
| Bihar | 2015 | All | 204,093 | 0.319 | Vital Statist. |
| Bihar | 2016 | All | 177,021 | 0.283 | Vital Statist. |
| Bihar | 2017 | All | 261,425 | 0.427 | Vital Statist. |
| Bihar | 2018 | All | 213,989 | 0.346 | Vital Statist. |
| Bihar | 2019 | All | 359,349 | 0.516 | Vital Statist. |
| Bihar | 2018 | Entire year | 182,921 |  | devdatalab |
|  | 2019 | Entire year | 351,274 |  | devdatalab |
|  | 2020 | Entire year | 387,429 |  | devdatalab |
| Bihar | 2018 | Jan-May | 45,487 |  | devdatalab |
|  | 2019 | Jan-May | 133,224 |  | devdatalab |
|  | 2020 | Jan-May | 138,693 |  | devdatalab |
|  | 2021 | Jan-May | 215,746 |  | devdatalab |
| Chhattisgarh | 2018 | Entire year | 15,690 |  | Online portal<br>of the state<br>CRS<br>(devdatalab) |
| Chhattisgarh | 2019 | Entire year | 15,607 |  |  |
| Chhattisgarh | 2020 | Entire year | 63,237 |  |  |
| Chhattisgarh | 2018 | Months 1-5 | 6,802 |  | Online portal<br>of the state<br>CRS<br>(devdatalab) |
| Chhattisgarh | 2019 | Months 1-5 | 5,623 |  |  |
| Chhattisgarh | 2020 | Months 1-5 | 17,497 |  |  |
| Chhattisgarh | 2021 | Months 1-5 | 106,832 |  |  |
| Chhattisgarh | 2015 | Entire year | 168,034 | 0.875 | Vital Statistics |
| Chhattisgarh | 2016 | Entire year | 182,985 | 0.952 | Vital Statistics |
| Chhattisgarh | 2017 | Entire year | 175,035 | 0.888 | Vital Statistics |
| Chhattisgarh | 2018 | Entire year | 177,549 | 0.835 | Vital Statistics |
| Chhattisgarh | 2019 | Entire year | 188,211 | 0.815 | Vital Statistics |
| Delhi | 2015 | All | 124,516 | 1.000 | Vital Statistics |
| Delhi | 2016 | All | 141,632 | 1.000 | Vital Statistics |
| Delhi | 2017 | All | 136,117 | 1.000 | Vital Statistics |
| Delhi | 2018 | All | 145,533 | 1.000 | Vital Statistics |
| Delhi | 2019 | All | 145,284 | 1.000 | Vital Statistics |
| Delhi | 2019 | Apr-May | 19,047 |  | Hindust. Tim. |
| Delhi | 2020 | Apr-May | 10,258 |  | Hindust. Tim. |
| Delhi | 2021 | Apr-May | 33,109 |  | Hindust. Tim. |
| Gujarat | 2015 | All | 412,322 | 1.000 | Vital Statistics |
| Gujarat | 2016 | All | 417,835 | 1.000 | Vital Statistics |
| Gujarat | 2017 | All | 388,316 | 0.982 | Vital Statistics |
| Gujarat | 2018 | All | 433,256 | 1.000 | Vital Statistics |
| Gujarat | 2019 | All | 462,284 | 1.000 | Vital Statistics |
| Gujarat | 2017 | Jan 1-Nov<br>30 | 368,000 |  | Dave |
| Gujarat | 2018 | Jan 1-Nov<br>30 | 393,000 |  | Dave |
| Gujarat | 2019 | Jan 1-Nov<br>30 | 419,000 |  | Dave |
| Gujarat | 2020 | Jan 1-Nov<br>30 | 374,000 |  | Dave |
| Gujarat | 2020 | Mar. 1-May<br>10 | 58,000 |  | Desai |
| Gujarat | 2021 | Mar. 1-May<br>10 | 123,871 |  | Desai |
| Gujarat:<br>Ahmedabad | 2017 | Mar. 1-May<br>10 | 9,319 |  | Opindia |
|  | 2018 | Mar. 1-May 10 | 9,866 |  | Opindia |
|  | 2019 | Mar. 1-May 10 | 9,950 |  | Opindia |
|  | 2020 | Mar. 1-May 10 | 7,786 |  | Opindia |
|  | 2021 | Mar. 1-May 10 | 13,593 |  | Opindia |
| Gujarat: Ahmedabad | 2019 | Apr-May | 5,490 |  | Khanna |
|  | 2020 | Apr-May | 10,708 |  | Khanna |
| Haryana | 2015 | Entire year | 168,910 | 1.000 | Vital Statistics |
|  | 2016 | Entire year | 181,138 | 1.000 | Vital Statistics |
|  | 2017 | Entire year | 174,937 | 1.000 | Vital Statistics |
|  | 2018 | Entire year | 185,842 | 1.000 | Vital Statistics |
|  | 2019 | Entire year | 188,910 | 1.000 | Vital Statistics |
| Haryana | 2018 | Entire year | 178,536 |  | Ramani |
|  | 2019 | Entire year | 184,155 |  | Ramani |
|  | 2020 | Entire year | 198,223 |  | Ramani |
| Haryana | 2018 | Jan-May | 74,816 |  | Ramani |
|  | 2019 | Jan-May | 76,870 |  | Ramani |
|  | 2020 | Jan-May | 78,030 |  | Ramani |
|  | 2021 | Jan-May | 121,100 |  | Ramani |
| Himachal Pradesh | 2015 | Entire year | 41,462 | 0.882 | Vital statistics |
|  | 2016 | Entire year | 35,819 | 0.734 | Vital statistics |
|  | 2017 | Entire year | 39,114 | 0.821 | Vital statistics |
|  | 2018 | Entire year | 41,833 | 0.834 | Vital statistics |
|  | 2019 | Entire year | 43,633 | 0.864 | Vital statistics |
| Himachal Pradesh | 2018 | Entire year | 42,151 |  | Ramani |
|  | 2019 | Entire year | 42,989 |  | Ramani |
|  | 2020 | Entire year | 44,436 |  | Ramani |
| Himachal Pradesh | 2018 | Jan-May | 17,759 |  | Ramani |
|  | 2019 | Jan-May | 17,515 |  | Ramani |
|  | 2020 | Jan-May | 16,806 |  | Ramani |
|  | 2021 | Jan-May | 21,180 |  | Ramani |
| Karnataka: Bengaluru | 2019 | Jan-July | 37,001 |  | The Hindu |
|  | 2020 | Jan-July | 49,135 |  | The Hindu |
|  | 2021 | Jan 1-June 15 | 87,082 |  | Chatterjee |
| Karnataka | 2015 | All | 393,731 | 0.962 | Srivat., Vital Statistics, devdatalab (all agree) |
| Karnataka | 2016 | All | 420,774 | 1.000 |  |
| Karnataka | 2017 | All | 481,747 | 1.000 |  |
| Karnataka | 2018 | All | 483,511 | 1.000 |  |
| Karnataka | 2019 | All | 508,584 | 1.000 |  |
| Karnataka | 2020 | All | 551,808 |  | Srivatsa, devdatalab |
| Karnataka | 2018 | Jan-June 15 | 224,000 |  | Chatterjee |
| Karnataka | 2019 | Jan-June 15 | 235,000 |  | Chatterjee |
| Karnataka | 2021 | Jan-June 15 | 337,580 |  | Chatterjee |
| Karnataka | 2015 | Jan-June | 194,466 |  | devdatalab |
| Karnataka | 2016 | Jan-June | 203,532 |  | devdatalab |
| Karnataka | 2017 | Jan-June | 229,367 |  | devdatalab |
| Karnataka | 2018 | Jan-June | 244,399 |  | devdatalab |
| Karnataka | 2019 | Jan-June | 235,151 |  | devdatalab |
| Karnataka | 2020 | Jan-June | 225,304 |  | devdatalab |
| Karnataka | 2021 | Jan 1-June 30 | 364,071 |  | karnataka.gov |
| Kerala | 2015 | Jan 1-May 31 | 95,281 |  | devdatalab |
| Kerala | 2016 | Jan 1-May 31 | 98,195 |  | devdatalab |
| Kerala | 2017 | Jan 1-May 31 | 96,534 |  | devdatalab |
| Kerala | 2018 | Jan 1-May 31 | 99,473 |  | devdatalab |
| Kerala | 2019 | Jan 1-May 31 | 104,705 |  | devdatalab |
| Kerala | 2020 | Jan 1-May 31 | 95,544 |  | devdatalab |
| Kerala | 2021 | Jan 1-May 31 | 113,372 |  | devdatalab |
| Kerala | 2015 | Entire year | 235,982 |  | Rajendran |
| Kerala | 2016 | Entire year | 244,900 |  | Rajendran |
| Kerala | 2017 | Entire year | 252,103 |  | Rajendran |
| Kerala | 2018 | Entire year | 255,594 |  | Rajendran |
| Kerala | 2019 | Entire year | 264,150 |  | Rajendran |
| Kerala | 2020 | Entire year | 252,421 |  | Rajendran |
| Kerala | 2021 | Jan 1-May 31 | 113,372 |  | Rajendran |
| Kerala | 2015 | All | 252,576 | 1.000 | Vital Statistics |
| Kerala | 2016 | All | 256,130 | 0.943 | Vital Statistics |
| Kerala | 2017 | All | 263,342 | 1.000 | Vital Statistics |
| Kerala | 2018 | All | 258,530 | 1.000 | Vital Statistics |
| Kerala | 2019 | All | 270,567 | 1.000 | Vital Statistics |
| Kerala | 2015 | All | 236,859 |  | lsgkerala.gov |
| Kerala | 2016 | All | 244,894 |  | lsgkerala.gov |
| Kerala | 2017 | All | 252,097 |  | lsgkerala.gov |
| Kerala | 2018 | All | 255,571 |  | lsgkerala.gov |
| Kerala | 2019 | All | 264,131 |  | lsgkerala.gov |
| Kerala | 2020 | All | 242,910 |  | lsgkerala.gov |
| Kerala | 2021 | Jan 1-July 1 | 115,778 |  | lsgkerala.gov |
| Madhya Pradesh | 2015 | All | 311,411 | 0.538 | Vital Statistics |
| Madhya Pradesh | 2016 | All | 338,587 | 0.609 | Vital Statistics |
| Madhya Pradesh | 2017 | All | 370,538 | 0.687 | Vital Statistics |
| Madhya Pradesh | 2018 | All | 424,527 | 0.788 | Vital Statistics |
| Madhya Pradesh | 2019 | All | 493,328 | 0.891 | Vital Statistics |
| Madhya Pradesh | 2018 | Months 1-12 | 407,172 |  | LM |
| Madhya Pradesh | 2019 | Months 1-12 | 449,819 |  | LM |
| Madhya Pradesh | 2020 | Months 1-12 | 461,057 |  | LM |
| Madhya Pradesh | 2018 | Months 1-5 | 152,346 |  | LM |
| Madhya Pradesh | 2019 | Months 1-5 | 167,549 |  | LM |
| Madhya Pradesh | 2020 | Months 1-5 | 164,191 |  | LM |
| Madhya Pradesh | 2021 | Months 1-5 | 348,708 |  | LM |
| Madhya Pradesh: urban regions of 25 districts | 2015 | Entire year | 119,068 |  | Vital Statistics |
|  | 2016 | Entire year | 112,303 |  | Vital Statistics |
|  | 2017 | Entire year | 113,612 |  | Vital Statistics |
|  | 2018 | Entire year | 112,188 |  | Vital Statistics |
|  | 2019 | Entire year | 119,914 |  | Vital Statistics |
|  | 2021 | April 1-30 | 21,456 |  | Datta |
| Maharashtra | 2015 | Entire year | 673,824 | 0.975 | Vital Statistics |
| Maharashtra | 2016 | Entire year | 666,448 | 0.937 | Vital Statistics |
| Maharashtra | 2017 | Entire year | 647,161 | 0.931 | Vital Statistics |
| Maharashtra | 2018 | Entire year | 667,900 | 0.984 | Vital Statistics |
| Maharashtra | 2019 | Entire year | 693,800 | 1.000 | Vital Statistics |
| Maharashtra | 2018 | Entire year | 426,489 |  | Ramani |
| Maharashtra | 2019 | Entire year | 462,028 |  | Ramani |
| Maharashtra | 2020 | Entire year | 565,180 |  | Ramani |
| Maharashtra | 2018 | Jan-May | 177,130 |  | Ramani |
| Maharashtra | 2019 | Jan-May | 186,004 |  | Ramani |
| Maharashtra | 2020 | Jan-May | 197,676 |  | Ramani |
| Maharashtra | 2021 | Jan-May | 287,123 |  | Ramani |
| Maharashtra:<br>Mumbai City | 2017 | Entire year | 89,037 |  | WM |
|  | 2018 | Entire year | 88,852 |  | WM |
|  | 2019 | Entire year | 91,123 |  | WM |
|  | 2020 | Entire year | 109,398 |  | WM |
| Maharashtra:<br>Mumbai City | 2017 | Jan-May | 36,166 |  | WM |
|  | 2018 | Jan-May | 37,173 |  | WM |
|  | 2019 | Jan-May | 37,363 |  | WM |
|  | 2020 | Jan-May | 35,914 |  | WM |
|  | 2021 | Jan-May | 45,163 |  | WM |
| Maharashtra:<br>Nagpur City | 2019 | Months 4-12 | 16,238 |  | WM |
|  | 2020 | Months 4-12 | 20,382 |  | WM |
| Odisha | 2015 | All year | 321,009 | 1.000 | Vital statistics |
| Odisha | 2016 | All year | 345,527 | 1.000 | Vital statistics |
| Odisha | 2017 | All year | 322,660 | 1.000 | Vital statistics |
| Odisha | 2018 | All year | 328,799 | 1.000 | Vital statistics |
| Odisha | 2019 | All year | 342,947 | 1.000 | Vital statistics |
| Odisha | 2021 | Jan 1-June 30 | 207,185 |  | odisha.gov.in |
| Punjab | 2015 | Entire year | 199,461 | 1.000 | Vital statistics |
| Punjab | 2016 | Entire year | 213,578 | 1.000 | Vital statistics |
| Punjab | 2017 | Entire year | 210,398 | 1.000 | Vital statistics |
| Punjab | 2018 | Entire year | 213,234 | 1.000 | Vital statistics |
| Punjab | 2019 | Entire year | 215,045 | 1.000 | Vital statistics |
| Punjab | 2015 | Entire year | 199,749 |  | Ramani |
| Punjab | 2016 | Entire year | 210,848 |  | Ramani |
| Punjab | 2017 | Entire year | 208,005 |  | Ramani |
| Punjab | 2018 | Entire year | 211,213 |  | Ramani |
| Punjab | 2019 | Entire year | 213,122 |  | Ramani |
| Punjab | 2020 | Entire year | 228,136 |  | Ramani |
| Punjab | 2015 | Jan-June | 100,258 |  | Ramani |
| Punjab | 2016 | Jan-June | 101,885 |  | Ramani |
| Punjab | 2017 | Jan-June | 101,688 |  | Ramani |
| Punjab | 2018 | Jan-June | 105,856 |  | Ramani |
| Punjab | 2019 | Jan-June | 107,823 |  | Ramani |
| Punjab | 2020 | Jan-June | 106,371 |  | Ramani |
| Punjab | 2021 | Jan-June | 147,389 |  | Ramani |
| Rajasthan | 2015 | Entire year | 409,463 | 0.899 | Vital statistics |
| Rajasthan | 2016 | Entire year | 416,992 | 0.933 | Vital statistics |
| Rajasthan | 2017 | Entire year | 424,763 | 0.953 | Vital statistics |
| Rajasthan | 2018 | Entire year | 443,173 | 0.999 | Vital statistics |
| Rajasthan | 2019 | Entire year | 451,315 | 0.986 | Vital statistics |
| Rajasthan | 2018 | Entire year | 216,370 |  | Ramani |
| Rajasthan | 2019 | Entire year | 219,814 |  | Ramani |
| Rajasthan | 2020 | Entire year | 229,564 |  | Ramani |
| Rajasthan | 2018 | Months 1-5 | 95,484 |  | Ramani |
| Rajasthan | 2019 | Months 1-5 | 90,366 |  | Ramani |
| Rajasthan | 2020 | Months 1-5 | 94,566 |  | Ramani |
| Rajasthan | 2021 | Months 1-5 | 125,863 |  | Ramani |
| Tamil Nadu, 6 cities: Madurai, Coimbatore, Trichy, Vellore, Karur, Tirupur. | 2019 | Jan-May | 10,587 |  | Nagar |
|  | 2020 | Jan-May | 9,432 |  | Nagar |
|  | 2021 | Jan-May | 18,587 |  | Nagar |
| Tamil Nadu: Madurai District. | 2019 | All | 19,735 |  | Radhakrishnan |
|  | 2020 | All | 21,524 |  | Radhakrishnan |
|  | 2019 | Jan-May | 8,789 |  | Radhakrishnan |
|  | 2020 | Jan-May | 7,724 |  | Radhakrishnan |
|  | 2021 | Jan-May | 11,208 |  | Radhakrishnan |
| Tamil Nadu | 2015 | All year | 568,271 | 1.000 | Vital Statistics |
|  | 2016 | All year | 563,625 | 1.000 | Vital Statistics |
|  | 2017 | All year | 580,496 | 1.000 | Vital Statistics |
|  | 2018 | All year | 574,006 | 1.000 | Vital Statistics |
|  | 2019 | All year | 633,897 | 1.000 | Vital Statistics |
| Tamil Nadu | 2018 | Entire year | 549,209 |  | crstn.org |
|  | 2019 | Entire year | 637,270 |  | crstn.org |
|  | 2020 | Entire year | 687,488 |  | crstn.org |
|  | 2021 | Jan 1-June 30 | 469,256 |  | crstn.org |
| Tamil Nadu | 2018 | All year | 536,192 |  | LM; Ramani |
|  | 2019 | All year | 588,221 |  | LM; Ramani |
|  | 2020 | All year | 644,291 |  | LM; Ramani |
|  | 2018 | Jan-May | 236,838 |  | LM; Ramani |
|  | 2019 | Jan-May | 250,289 |  | LM; Ramani |
|  | 2020 | Jan-May | 245,328 |  | LM; Ramani |
|  | 2021 | Jan-May | 318,245 |  | LM; Ramani |
| Tamil Nadu: Chennai | 2015 | Weeks 1-52 | 59,875 |  | LM |
|  | 2016 | Weeks 1-52 | 57,826 |  | LM |
|  | 2017 | Weeks 1-52 | 63,726 |  | LM |
|  | 2018 | Weeks 1-52 | 62,793 |  | LM |
|  | 2019 | Weeks 1-52 | 67,002 |  | LM |
|  | 2020 | Weeks 1-52 | 73,932 |  | LM |
|  | 2015 | Weeks 1-19 | 21,790 |  | LM |
|  | 2016 | Weeks 1-19 | 21,544 |  | LM |
|  | 2017 | Weeks 1-19 | 22,495 |  | LM |
|  | 2018 | Weeks 1-19 | 23,002 |  | LM |
|  | 2019 | Weeks 1-19 | 23,912 |  | LM |
|  | 2020 | Weeks 1-19 | 22,122 |  | LM |
|  | 2021 | Weeks 1-19 | 35,854 |  | LM |
| Tamil Nadu:<br>Chennai | 2015 | Jan 1-June<br>30 | 29,450 |  | Rukmini |
|  | 2016 | Jan 1-June<br>30 | 29,477 |  | Rukmini |
|  | 2017 | Jan 1-June<br>30 | 31,131 |  | Rukmini |
|  | 2018 | Jan 1-June<br>30 | 31,140 |  | Rukmini |
|  | 2019 | Jan 1-June<br>30 | 33,391 |  | Rukmini |
|  | 2020 | Jan 1-June<br>30 | 34,579 |  | Rukmini |
|  | 2021 | Jan 1-June<br>30 | 54,748 |  | Rukmini |
| Telangana:<br>Hyderabad | 2016 | Jan-Dec | 49,523 |  | LM, Ramani |
|  | 2017 | Jan-Dec | 52,710 |  | LM, Ramani |
|  | 2018 | Jan-Dec | 55,026 |  | LM, Ramani |
|  | 2019 | Jan-Dec | 66,131 |  | LM, Ramani |
|  | 2020 | Jan-Dec | 77,241 |  | LM, Ramani |
| Telangana:<br>Hyderabad | 2016 | Jan-May | 18,839 |  | LM, Ramani |
|  | 2017 | Jan-May | 20,645 |  | LM, Ramani |
|  | 2018 | Jan-May | 21,696 |  | LM, Ramani |
|  | 2019 | Jan-May | 25,657 |  | LM, Ramani |
|  | 2020 | Jan-May | 24,884 |  | LM, Ramani |
|  | 2021 | Jan-May | 36,041 |  | LM, Ramani |
| Uttar Pradesh | 2015 | Entire year | 687,416 | 0.442 | Vital Statistics |
| Uttar Pradesh | 2016 | Entire year | 608,740 | 0.402 | Vital Statistics |
| Uttar Pradesh | 2017 | Entire year | 571,170 | 0.383 | Vital Statistics |
| Uttar Pradesh | 2018 | Entire year | 906,653 | 0.608 | Vital Statistics |
| Uttar Pradesh | 2019 | Entire year | 944,596 | 0.633 | Vital Statistics |
| Uttar Pradesh | 2019 | Entire year | 773,402 |  | Bhawan |
| Uttar Pradesh | 2020 | Entire year | 793,505 |  | Bhawan |
| Uttar Pradesh | 2019 | Jan 1-Apr<br>30 | 259,316 |  | Bhawan |
| Uttar Pradesh | 2020 | Jan 1-Apr<br>30 | 188,747 |  | Bhawan |
| Uttar Pradesh | 2021 | Jan 1-Apr<br>30 | 333,878 |  | Bhawan |
| West Bengal | 2015 | Entire year | 403,180 | 0.723 | Vital statistics |
| West Bengal | 2016 | Entire year | 445,540 | 0.806 | Vital statistics |
| West Bengal | 2017 | Entire year | 442,995 | 0.797 | Vital statistics |
| West Bengal | 2018 | Entire year | 490,530 | 0.908 | Vital statistics |
| West Bengal | 2019 | Entire year | 551,695 | 1.000 | Vital statistics |
| West Bengal | 2018 | Entire year | 368,477 |  | Ramani |
| West Bengal | 2019 | Entire year | 410,503 |  | Ramani |
| West Bengal | 2020 | Entire year | 463,858 |  | Ramani |
| West Bengal | 2018 | Jan-May | 155,108 |  | Ramani |
| West Bengal | 2019 | Jan-May | 167,422 |  | Ramani |
| West Bengal | 2020 | Jan-May | 176,451 |  | Ramani |
| West Bengal | 2021 | Jan-May | 216,972 |  | Ramani |
| West Bengal:<br>Kolkata | 2015 | Weeks 1-52 | 62,710 |  | LM |
|  | 2016 | Weeks 1-52 | 65,060 |  | LM |
|  | 2017 | Weeks 1-52 | 69,910 |  | LM |
|  | 2018 | Weeks 1-52 | 68,998 |  | LM |
|  | 2019 | Weeks 1-52 | 69,844 |  | LM |
|  | 2020 | Weeks 1-52 | 74,841 |  | LM |
| West Bengal:<br>Kolkata | 2015 | Weeks 1-20 | 25,128 |  | LM |
|  | 2016 | Weeks 1-20 | 24,506 |  | LM |
|  | 2017 | Weeks 1-20 | 26,980 |  | LM |
|  | 2018 | Weeks 1-20 | 25,985 |  | LM |
|  | 2019 | Weeks 1-20 | 29,222 |  | LM |
|  | 2020 | Weeks 1-20 | 27,674 |  | LM |
|  | 2021 | Weeks 1-20 | 33,132 |  | LM |

LM = Local Mortality dataset (Karlinsky 2021).
The urban areas in Madhya Pradesh from which funeral counts were tabulated for April 2021 were from the following 25 districts: Barwani, Bhind, Bhopal, Burhanpur, Chhatarpur, Chhindwara, Dewas, Dhar, Gwalior, Indore, Jabalpur, Jhabua, Khandwa, Mandsaur, Morena, Neemuch, Ratlam, Sagar, Satna, Seoni, Shahdol, Shivpuri, Singroli, Tikamgarh, Vidisha (Datta 2021).

## Notes

The author has no conflicts of interest to disclose.

### Competing Interest Statement

The authors have declared no competing interest.

### Funding Statement

No funding.

### Author Declarations

This study was approved by the Virginia Commonwealth University Office of Research Subjects Protection.

## References

Alavi AM. NDTV. Bihar Saw Nearly 75,000 Unaccounted Deaths Amid 2nd Covid Wave, Data Shows. June 19, 2021. Available from: https://www.ndtv.com/india-news/bihar-saw-nearly-75-000-unaccounted-deaths-amid-2nd-covid-wave-data-shows-2467778?pfrom=home-ndtv_topscroll Accessed June 20, 2021.

Aldujeli A, Hamadeh A, Briedis K, Tecson KM, Rutland J, Krivickas Z, Stiklioraitis S, et al. Delays in presentation in patients with acute myocardial infarction during the COVID-19 pandemic. Cardiology Research 11, no. 6 (2020): 386.

Anand A, Sandefur J, Subramanian A. Three new estimates of India’s all-cause excess mortality during the COVID-19 pandemic. Center for Global Development. July 20, 2021. Available from: https://cgdev.org/publication/three-new-estimates-indias-all-cause-excess-mortality-during-covid-19-pandemic Accessed July 25, 2021.

Anparthi A. 128 more deaths than normal ones, Covid fatalities in Dec ‘20 [in Nagpur]. Times of India. January 21, 2021. Available from: https://timesofindia.indiatimes.com/city/nagpur/128-more-deaths-than-normal-ones-covid-fatalities-in-dec-20/articleshow/80373223.cms Accessed June 16, 2021.

Bhawan S. Deputy Chief Registrar. Vital Dept (Birth & Death). Government of Uttar Pradesh. Information furbished for RTI [requested by Saurav Das]. June 2, 2021.

Chatterjee S. Karnataka recorded 1.02 lakh ‘excess’ deaths in 2021, 5 times the COVID-19 toll. The News Minute. June 15, 2021. Available from: https://www.thenewsminute.com/article/karnataka-recorded-102-lakh-excess-deaths-2021-5-times-covid-19-toll-150761 Accessed June 16, 2021.

Das S. Death Count In 24 UP Districts 43 Times More Than Official Covid-19 Toll. Article14. June 21, 2021. Available from: https://article-14.com/post/untitled-60cf605395758 Accessed June 21, 2021.

Datta S. Madhya Pradesh Excess Deaths in April: An Alternative Count Using Funeral Data. Thewire.in. June 22, 2021. Available from: https://science.thewire.in/health/madhya-pradesh-excess-deaths-covid-19-april-2021-funeral-data/ Accessed June 26, 2021.

Dave K. Year of pandemic? Gujarat records 11% less deaths than in 2019. Times of India. December 13, 2020. Available from: https://timesofindia.indiatimes.com/city/ahmedabad/year-of-pandemic-gujarat-records-11-less-deaths-than-in-2019/articleshow/79704719.cms Accessed June 19, 2021.

Debroy S. 13,000 more deaths in Mumbai this year between March & September. Times of India. October 29, 2020. Available from: https://timesofindia.indiatimes.com/city/mumbai/13k-more-deaths-in-city-this-year-between-march-sept/articleshow/78920631.cms Accessed June 27, 2021.

Desai D. 61k Covid deaths not counted in Gujarat: Report. Hindustan Times. May 15, 2021. Available from: https://www.hindustantimes.com/india-news/61k-covid-deaths-not-counted-in-gujarat-report-101621027267608.html Accessed June 14, 2021.

Deshmukh Y, Suraweera W, Tumbe C, Bhowmick A, Sharma S, Novosad P, Fu SH, Newcombe L, Gelband H, Brown P, Jha P. Excess mortality in India from June 2020 to June 2021 during the COVID pandemic: death registration, health facility deaths, and survey data. Medrxiv. July 23, 2021. Available from: https://www.medrxiv.org/content/10.1101/2021.07.20.21260872v1 Accessed July 25, 2021.

Development Data Lab. Making Sense of Excess Mortality. July 1, 2021. Available from: https://devdatalab.medium.com/ Accessed July 23, 2021.

Government of India. Office of the Registrar General. Ministry of Home Affaris. Vital Statistics of India. Based on the Civil Registration System. 2019. June 2021. Available from: https://crsorgi.gov.in/web/uploads/download/CRS%202019%20report.pdf Accessed June 18, 2021.

Government of India. Office of the Registrar General. Ministry of Home Affaris. Vital Statistics of India. Based on the Civil Registration System. 2018. June 2020.

https://censusindia.gov.in/2011-Common/CRS_2018/crs2018_20072020.pdf Accessed June 18, 2021.

Government of India. Office of the Registrar General. Ministry of Home Affaris. Vital Statistics of India. Based on the Civil Registration System. 2015. June 2017. Available from: https://censusindia.gov.in/2011-Documents/CRS_Report/crs_report%202015_23062017.pdf Accessed June 18, 2021.

Government of India. Office of the Registrar General. Ministry of Home Affaris. Vital Statistics of India. Based on the Civil Registration System. 2016. June 2018. Available from: https://censusindia.gov.in/2011-Documents/CRS_Report/CRS%20FINAL%20REPORT%202016_21062018.pdf Accessed June 18, 2021.

Government of India. Office of the Registrar General. Ministry of Home Affaris. Vital Statistics of India. Based on the Civil Registration System. 2017. August 2019. Available from: https://censusindia.gov.in/2011-Documents/CRS_Report/CRS_report_2017_2020_02_26_revised.pdf Accessed June 18, 2021.

Government of Karnataka. Annual Report on the Registration of Births and Deaths Act, 1969. [2015-2019]. Available from: https://ejanma.karnataka.gov.in/frmVitalSat.aspx Accessed June 14, 2021.

Government of Karnataka. Office of the Chief Registrar of Births and Deaths. Registration Details. Available from: https://ejanma.karnataka.gov.in/frmTransaction_Details.aspx Accessed July 2, 2021

Government of Kerala. Civil Registrations. June 20, 2021. Available from: https://cr.lsgkerala.gov.in/Pages/map.php Accessed July 1, 2021.

Government of Odisha. Health and Family Welfare Department. Birth and Death Odisha. Available from: https://www.birthdeath.odisha.gov.in/#/home Accessed July 1, 2021.

Government of Tamil Nadu. Birth and Death Registration. June 27, 2021 [in India]. Available from: http://www.crstn.org/birth_death_tn/MisRep.jsp Accessed June 26, 2021.

Guilmoto CZ. Estimating the death toll of the Covid-19 pandemic in India. Medrxiv. July 2, 2021. Available from: https://www.medrxiv.org/content/10.1101/2021.06.29.21257965v1 Accessed July 25, 2021.

[The Hindu] Special Correspondent. 32% increase in mortality rate in Bengaluru: Patil. The Hindu. Sep. 3, 2020. Available from: https://www.thehindu.com/news/national/karnataka/32-increase-in-mortality-rate-in-bengaluru-patil/article32509298.ece Accessed: June 16, 2021.

Hindustan Times Correspondent. What do Delhi’s death registration figures tell us about Covid-19 death toll? Hindustan Times. June 4, 2021. Available from: https://www.msn.com/en-in/news/other/what-do-delhi-s-death-registration-figures-tell-us-about-covid-19-death-toll/ar-AAKHLUM Accessed June 6, 2021.

IANS. COVID-19 year 2021 sees 1.4L more deaths than 2019 in Tamil Nadu, says a study by NGO [Arappor Iyakkam]. The Free Press Journal. June 26, 2021. Available from: https://www.freepressjournal.in/india/covid-19-year-2021-sees-14l-more-deaths-than-2019-in-tamil-nadu-says-a-study-by-ngo Accessed June 26, 2021.

Karlinsky A, et. al. Local Mortality Dataset. Available from: https://github.com/akarlinsky/world_mortality/tree/main/local_mortality Accessed June 6, 2021.

Karlinsky A, Kobak D. The World Mortality Dataset: Tracking excess mortality across countries during the COVID-19 pandemic. eLife 2021;10:e69336. Available from: https://elifesciences.org/articles/69336 Accessed July 25, 2021.

Johns Hopkins University. COVID-19 Data Repository by the Center for Systems Science and Engineering (CSSE) at Johns Hopkins University. Available from: https://github.com/CSSEGISandData/COVID-19 Accessed June 6, 2021.

IHME, University of Washington. Covid-19 Projections. India. Cumulative Deaths. July 25, 2021a. Available from: https://covid19.healthdata.org/india?view=cumulative-deaths&tab=trend Accessed July 25, 2021.

IHME, University of Washington. Estimation of total mortality due to COVID-19. May 13, 2021b. Available from: http://www.healthdata.org/special-analysis/estimation-excess-mortality-due-covid-19-and-scalars-reported-covid-19-deaths Accessed June 6, 2021.

Khanna S. Other deaths spike in Indian city [Ahmedabad] ravaged by coronavirus. Reuters. July 2, 2020. Available from: https://www.reuters.com/article/us-health-coronavirus-india-casualties-idUSKBN24311A Accessed June 16, 2021.

Nagar T. Citizen’s Research Report on Covid Deaths in TamilNadu. Report by Arappor Iyakkam. June 15, 2021. Available from: https://arappor.org/public/Document/20210615_Citizensreport_Coronadeath_withAnnexure.pdf Accessed June 16, 2021.

OpIndia Staff. Gujarat: Here’s why the death certificate stats from March to May so far do not prove allegations of massive undercounting of Covid deaths. OpIndia. May 15, 2021. Available from: https://www.opindia.com/2021/05/gujarat-covid-19-deaths-death-certificate-statistics-divya-bhaskar/ Accessed June 14, 2021. [Has Gujarat and Ahmedabad data.]

Radhakrishnan S. Is Madurai undercounting Covid deaths? Data raise questions. The New Indian Express. June 18, 2021. Available from: https://www.newindianexpress.com/states/tamil-nadu/2021/jun/18/is-madurai-undercounting-covid-deaths-data-raise-questions-2317785.html Accessed June 17, 2021.

Rajendran D, Kurian S. TNM Exclusive: Kerala reports around 14,000 excess deaths till end of May 2021. The News Minute. Available from: https://www.thenewsminute.com/article/tnm-exclusive-kerala-reports-almost-14000-excess-deaths-till-end-may-2021-151124 Accessed June 23, 2022.

Ramani S, Radhakrishnan V. Excess deaths in Hyderabad are 10 times the official COVID-19 toll for Telangana. The Hindu. June 14, 2021. Available from: https://www.thehindu.com/news/cities/Hyderabad/excess-deaths-in-hyderabad-are-10-times-the-official-covid-19-toll-for-telangana/article34807214.ece Accessed June 14, 2021.

Ramani S, Radhakrishnan V. Number of death certificates issued by GHMC in a month. Available from: https://docs.google.com/spreadsheets/d/1wP1SOEHclYOmA2hokjBzjCAO_eqpvx9LPFtxDOkv1zg/edit#gid=0 Accessed June 14, 2021.

Ramani S, Kannan R. Excess deaths in Tamil Nadu over four times official COVID-19 tally. The Hindu. June 16, 2021. https://www.thehindu.com/news/national/tamil-nadu/excess-deaths-in-tamil-nadu-over-four-times-official-covid-19-tally/article34834150.ece?homepage=true Accessed June 17, 2021.

Ramani S. Excess deaths in Rajasthan are at least five times the official COVID-19 tally. The Hindu. July 4, 2021. Available from: https://www.thehindu.com/news/national/excess-deaths-in-rajasthan-are-five-times-the-official-covid-19-tally/article35118826.ece Accessed July 23, 2021.

Ramani S. ‘Excess deaths’ in Haryana seven times official COVID-19 toll. The Hindu. July 14, 2021. Available from: https://www.thehindu.com/news/national/excess-deaths-in-haryana-seven-times-official-covid-19-toll/article35329023.ece Accessed July 24, 2021.

Ramani S, Vasudeva V. Punjab ‘excess deaths’ three times official toll. The Hindu. July 30, 2021. Available from: https://www.thehindu.com/news/national/coronavirus-punjab-excess-deaths-three-times-official-toll/article35641394.ece?homepage=true Accessed July 31, 2021.

Ramani S. Himachal Pradesh ‘excess deaths’ twice the official COVID-19 toll. The Hindu. July 21, 2021. Available from: https://www.thehindu.com/news/national/himachal-pradesh-excess-deaths-twice-the-official-covid-19-toll/article35430252.ece Accessed July 31, 2021.

Ramani S. Excess deaths in West Bengal 11 times official COVID-19 tally. The Hindu. July 25, 2021. Available from: https://www.thehindu.com/news/national/other-states/excess-deaths-in-west-bengal-11-times-official-covid-19-tally/article35526895.ece Accessed July 31, 2021.

Ramani S. Excess deaths in Maharashtra were at least 3 times the official COVID toll. The Hindu. August 3, 2021. Available from: https://www.thehindu.com/news/national/excess-deaths-in-maharashtra-were-at-least-3-times-the-official-covid-toll/article35708965.ece Accessed August 3, 2021.

Rukmini S. Madhya Pradesh saw nearly three times more deaths than normal after second wave of Covid-19 struck. Scroll.In. June 12, 2021. Available from: https://scroll.in/article/996772/madhya-pradesh-saw-nearly-three-times-more-deaths-than-normal-after-second-wave-of-covid-19-struck Accessed June 12, 2021.

Rukmini S. Andhra Pradesh saw 400% increase in deaths in May, Tamil Nadu saw more modest excess mortality. Scroll.In. June 13, 2021. Available from: https://scroll.in/article/997427/andhra-pradesh-saw-400-increase-in-deaths-in-may-tamil-nadu-saw-more-modest-excess-mortality Accessed June 13, 2021. [“AP”]

Rukmini S. How India Could Fill In The Blanks On Excess Mortality. India Spend. June 17, 2020. Available from: https://www.indiaspend.com/how-india-could-fill-in-the-blanks-on-excess-mortality/ Accessed June 27, 2021.

Rukmini S. Daily Registered Deaths in Chennai. Chennai Municipal Corporation. Available from: https://github.com/elseasama/covid19chennai/tree/gh-pages/chennai_data Accessed July 23, 2021.

Saikia A. Assam saw 28,000 more deaths than normal in months when first wave of Covid-19 struck. Scroll.in. June 16, 2021. Available from: https://scroll.in/article/997683/assam-saw-28000-more-deaths-than-normal-in-months-when-first-wave-of-covid-19-struck Accessed June 17, 2021.

Scroll Staff. Gujarat is undercounting Covid-19 deaths, shows ‘Divya Bhaskar’ report. Scroll.In. May 14, 2021. Available from: https://scroll.in/latest/994906/gujarat-is-undercounting-covid-19-deaths-shows-divya-bhaskar-report Accessed June 6, 2021.

Srivatsa SS, Ramani S. Karnataka’s excess deaths nearly 6 times official COVID-19 toll. The Hindu. June 20, 2021. Available from: https://www.thehindu.com/news/national/excess-deaths-in-karnataka-nearly-six-times-official-covid-19-tally/article34870624.ece Accessed June 21, 2021.

Srivatsa SS, Ramani S. Number of deaths registered in CRS system of Karnataka. June 21, 2021. Available from: https://docs.google.com/spreadsheets/d/1pBM4wFuGQgnL6syAG3IFHGUk9yzc4pToSv2tftKElxY/edit#gid=0 Accessed June 21, 2021.

Wu Y, Chen F, Wang Z, Feng W, Liu Y, Wang Y, Song H. Reductions in hospital admissions and delays in acute stroke care during the pandemic of COVID-19. Frontiers in Neurology 11 (2020): 1251.

